# Hospitalizations for Body Dysmorphic Disorder Across the U.S: Insights from the Nationwide Inpatient Sample Database

**DOI:** 10.1101/2024.08.17.24312149

**Authors:** Oluwatobi Olaomi, Kenechukwu Anona, Silvia Udegbe, Fidelis Uwumiro, Nnenna Okafor, Chinwendu Obijuru, Halleluya Yayehyirad, Mojeed Opeyemi, Micheal Bojerenu, Victory Okpujie, Nnaedozie Umeani

**Affiliations:** University of Ibadan, College of Medicine, Oyo State, Nigeria; Greater Manchester Mental Health NHS Foundation Trust, UK; Ambrose Alli University, Ekpoma, Edo State, Nigeria; Southern Regional Medical Center, Riverdale, Georgia, USA; All Saints University College of Medicine, Belair Kingstown, St. Vincent and the Grenadines; College of Medicine, University of Nigeria, Ituku-Ozalla, Enugu, Nigeria; Oxleas NHS Foundation Trust, London, UK; Federal Medical Center, Abeokuta, Ogun State, Nigeria; St. Barnabas Hospital SBH Health System, Bronx, NY, USA; University of Benin Teaching Hospital, Benin, Edo State, Nigeria; College of Medicine, University of Lagos, Lagos Nigeria

**Keywords:** Body dysmorphic disorder, dysmorphophobia, obsessive-compulsive disorders, suicidality, body image disorder, nationwide inpatient sample

## Abstract

**Background/Aim:** Body dysmorphic disorder (BDD) has been associated with significant psychiatric comorbidity and suicidal ideation in the outpatient setting. We examined the hospitalization rates, resource utilization, comorbidities, and outcomes of BDD using inpatient data. In addition, we compared the outcomes of BDD hospitalizations with those of patients with other mental disorders.

**Methods:** Adult hospitalizations for a primary diagnosis of BDD were identified from the 2017-2020 nationwide inpatient sample database using the ICD-10 code (F45.22). We used χ2 analysis to compare categorical variables. For continuous variables, we used t-tests and the Wilcoxon rank-sum test to analyze data with normal and nonnormal distributions. Using rank commands and ICD-10-CM/PCS codes, we identified the most recurring comorbidities and procedures associated with hospitalizations for BDD. The study endpoints included hospitalization rates, secondary indications for hospitalization, most recurring comorbidities and procedures, and outcomes (hospitalization costs, length of stay, and discharge disposition).

**Results:** 3,384 BDD hospitalizations were analyzed (67 cases per 100,000 hospitalizations for mental disorders). 67.6% were female, whereas non-BDD admissions had a male preponderance (67.5%). 71.4% of BDD hospitalizations were for individuals aged between 18 and 40 years. Hospitalizations for BDD were younger than those for other mental disorders (median age: 33 [IQR, 24-50] vs 37 [IQR, 24-54], P<0.001). Approximately 94.6% of BDD hospitalizations had at least one other psychiatric comorbidity. The most common comorbidities were social anxiety disorder (40%), major depressive disorder (22.8%), bipolar II disorder (2.2%), and substance use disorder (18.9%). 17 individuals (O.5%) had attempted suicide, while 10.3% received various cosmetic procedures. BDD was associated with a longer mean hospital stay compared with other mental disorders combined (9 vs 7.8 days; P=0.002). 89.8% of BDD hospitalizations were discharged home routinely, and no mortality was recorded over the period. BDD hospitalizations were associated with higher mean hospital costs than hospitalizations for other mental disorders combined ($34,314 vs. $27,770; P<0.001).

**Conclusion:** BDD hospitalizations are most common among young women, and involve significant psychiatric comorbidities, longer hospital stays, and higher costs than other mental health conditions.

## INTRODUCTION

Body dysmorphic disorder (BDD) or dysmorphophobia, classified under the category of obsessive-compulsive and other related disorders in the DSM-5, is a psychiatric condition marked by an intense fixation on perceived imperfections or defects in one’s physical appearance, which might be unnoticeable or slightly discernible to others.^1,2^ Historical accounts trace its description back to Emil Kraepelin and Pierre Janet in the 1800s. Recent studies place its prevalence at approximately 2%–3% in the general population, with some reports indicating higher incidence rates in cosmetic surgery contexts.^2,3^ Indeed, those with BDD may frequently compare their features with others, seeking constant reassurance and exhibiting signs of diminished self-esteem. The pathogenesis of BDD remains somewhat elusive, although it has been postulated that its origins stem from a combination of biological, psychosocial, and sociocultural factors.^4^ Many symptoms manifest early in life and are potentially triggered by traumatic experiences or abuse that hinder normal psychological development, consequently affecting the capacity for meaningful interpersonal relationships.^4^ Both genders exhibit unique areas of bodily concern: women might be preoccupied with aspects such as skin, hair, or nose, whereas men might focus on chin, teeth, or body build.^5^

Comorbidities with other psychiatric conditions are common in patients with BDD. Major depressive disorder, anxiety disorder, substance abuse, and obsessive-compulsive disorder have all been observed in individuals with this disorder in various clinical settings.^6,7,8^ A considerable association between BDD and suicidal ideation has been identified. For instance, Phillips and Menard reported that 57.8% of patients with BDD had suicidal thoughts and 2.6% had attempted suicide.^9^ Patients with BDD often approach dermatological or cosmetic clinics, possibly because of a lack of awareness about BDD or available treatments, such as antidepressants and cognitive behavioral therapy, or because of fears of discussing their perceived flaws with mental health professionals.^10,11^ Despite the numerous interventions they may pursue, many patients experience little to no improvement or even worsening of their BDD symptoms after undergoing cosmetic procedures, exacerbating their negative cognition and suicide risk.^12^

Despite the growing literature linking BDD to other psychiatric conditions and cosmetic procedures, hospitalization rates and outcomes in patients with BDD remain understudied. This index study explores the hospitalization rates, characteristics, comorbidities, and outcomes among patients with BDD.

## METHODS

### Data source

This study used data from the Nationwide Inpatient Sample (NIS) database, created by the Agency for Healthcare Research and Quality (AHRQ) as part of the Healthcare Cost and Utilization Project (HCUP). As the largest publicly accessible inpatient database in the United States, the NIS has recorded data from approximately 8 million hospital stays annually across approximately 4,000 hospitals since 2012. When weighted, the NIS data represents approximately 35 million hospitalizations nationwide each year. The database adopts a stratified sampling approach for hospital selection, considering variables such as geographical location, teaching status, and hospital bed size. This method yields comprehensive insights into healthcare utilization, including length of stay, total costs, and patient outcomes. The NIS meticulously compiles up to 40 distinct diagnoses and 15 procedures per hospital stay, with a strict policy of anonymization of every record entry. The primary diagnosis is the main reason for the hospitalization. The secondary diagnosis includes all other diagnoses. The NIS encompasses diverse information, including patient demographics, comorbidities, procedures performed, insurance details, total hospital charges, and in-hospital outcomes, enabling detailed multiyear studies.^13,14,15^

### Selection criteria and study endpoints

We queried the NIS database to identify all adult hospitalizations for mental diseases and disorders using the major diagnostic categories (MDC-19). From this cohort, we identified hospitalizations for BDD using the ICD-10-CM code (F45.22). The outcomes of interest included hospitalization rates, indications for hospitalization, and the most recurring comorbidities and procedures. In addition, we compared the sociodemographic characteristics and hospitalization outcomes (including discharge disposition, mean length of hospital stay, and hospitalization costs) between patients with BDD and those with other mental diseases and disorders.

### Statistical analysis

In compliance with the recommendations of the AHRQ, weighted data were used for all statistical analyses.^16^ We accounted for clustering (HOSP_NIS), weighting (DISCWT), and stratification (NIS_STRATUM) within the Nationwide Inpatient Sample during this study, ensuring the use of only population-representative data. First, we compared the sociodemographic characteristics of BDD hospitalizations, including patient- and hospital-level variables, with those of other hospitalization for mental diseases and disorders. We used Pearson’s χ2 analysis or Fisher’s exact test to compare categorical variables. For continuous variables, we used t-tests and the Wilcoxon rank-sum test to analyze data with normal and nonnormal distributions. The normality of the data distribution was assessed for the age variable using graphical methods and by skewness and kurtosis using the Kolmogorov–Smirnov test for normality of distributions. Using rank commands and ICD-10-CM/PCS codes, we identified the most recurring comorbidities and procedures associated with hospitalizations for BDD.

**Figure 1:**
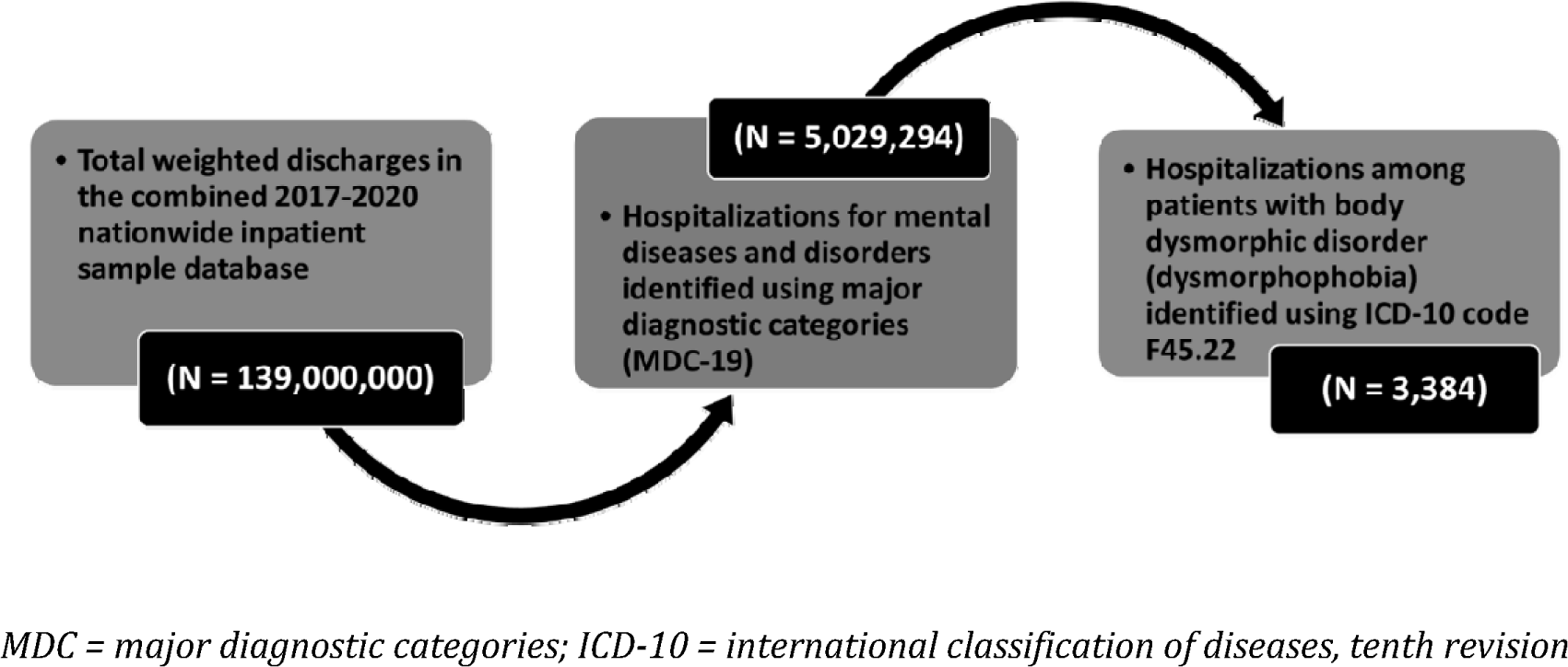
Selection criteria.

## RESULTS

### Sociodemographic characteristics and indications for hospitalization

A total of 3,384 BDD hospitalizations were identified from a cohort of 5,029,294 adult hospitalizations for mental diseases and disorders (amounting to 67 cases of BDD per 100,000 hospitalizations for mental diseases or disorders). The median age of individuals with BDD was 33 (IQR 24-50) years, which was lower than that of individuals with other mental disorders (37; IQR, 24-54; P<0.001). Gender distribution differed notably between the BDD cohort and non-BDD admissions. Among BDD admissions, 67.6% were female, whereas non-BDD admissions had a male preponderance (67.5%). Additionally, most BDD hospitalizations (71.4%) occurred in individuals aged between 18 and 40 years, in contrast to other admissions (Table 1). Regarding admission characteristics, most BDD admissions (88.5%) were nonelective and occurred on weekdays (79.9%). Furthermore, these admissions were concentrated primarily in large urban teaching hospitals (73.6%).

**Table 1:** Baseline sociodemographic characteristics and resource use of BDD and non-BDD hospitalizations.

| **Patient and hospital variables** | **Non-BDD hospitalizations**<br>**(N = 5,025,910)** | **BDD hospitalizations**<br>**(N = 3,384)** | **P-value\*** |
| --- | --- | --- | --- |
| Female sex, % | 32.5 | 67.6 | <0.001 |
| Median age, years (IQR) | 37 (24-54) | 34 (24-50) | <0.001 |
| Age categories, years |  |  | <0.001 |
| Age ≥ 18 and 40 | 48.6 | 71.4 |  |
| Age ≥ 41 and 60 | 33.4 | 22.8 |  |
| > 60 | 18 | 5.8 |  |
| Elective admissions, % | 12.6 | 11.5 | 0.535 |
| Race/ethnicity, % |  |  | <0.001 |
| White | 63.5 | 75.6 |  |
| Black | 20.8 | 7.0 |  |
| Hispanic | 10.0 | 8.1 |  |
| Asian or Pacific Islander | 1.7 | 4.7 |  |
| Native American | 0.6 | 0.3 |  |
| Other | 3.3 | 4.4 |  |
| Admission day is a weekend, % | 20.3 | 20.1 | 0.899 |
| Charlson comorbidity index score, % |  |  | <0.001 |
| 0 | 63.9 | 83.8 |  |
| 1 | 22.0 | 12.9 |  |
| 2 | 7.5 | 2.8 |  |
| ≥ 3 | 6.6 | 0.6 |  |
| Comorbidities, % |  |  |  |
| Acute myocardial infarction | 1.8 | 0.8 | 0.162 |
| Congestive heart failure | 2.9 | 0.8 | 0.018 |
| Peripheral vascular disease | 1.2 | 0.3 | 0.118 |
| Cerebrovascular disease | 1.4 | 0.3 | 0.691 |
| Dementia | 7.2 | 1.7 | <0.001 |
| COPD | 16.7 | 8.2 | <0.001 |
| Rheumatoid disease | 1.1 | 1.1 | 0.988 |
| Peptic ulcer disease | 0.3 | 0.3 | 0.904 |
| Mild liver disease | 2.1 | 0.8 | 0.096 |
| Diabetes uncomplicated | 10.9 | 2.5 | <0.001 |
| Diabetes + complications | 2.8 | 0.8 | 0.021 |
| Hemiplegia or paraplegia | 0.4 | 0.0 | 0.288 |
| Renal disease | 3.6 | 1.4 | 0.023 |
| Cancer | 0.7 | 0.0 | 0.153 |
| Moderate/severe liver disease | 0.2 | 0.0 | 0.482 |
| Metastatic cancer | 0.2 | 0.0 | 0.494 |
| AIDS | 0.5 | 0.0 | 0.246 |
| Median household income (quartiles) ** |  |  | <0.001 |
| First (0-25th) | 36.4 | 24.9 |  |
| Second (26th– 50th) | 26.8 | 22.1 |  |
| Third (51st-75<sup>th</sup>) | 21.0 | 21.6 |  |
| Fourth (76th-100th) | 15.8 | 31.4 |  |
| Insurance type, % |  |  | <0.001 |
| Medicare | 32.1 | 20.2 |  |
| Medicaid | 36.5 | 22.4 |  |
| Private | 24.2 | 52.0 |  |
| Self-pay | 7.3 | 5.4 |  |
| Hospital region, % |  |  | <0.001 |
| Northeast | 22.0 | 28.6 |  |
| Midwest | 26.1 | 23.4 |  |
| South | 36.1 | 27.2 |  |
| West | 15.7 | 20.9 |  |
| Hospital bed size, % |  |  | 0.919 |
| Small | 23.5 | 22.5 |  |
| Medium | 28.1 | 28.6 |  |
| Large | 48.4 | 48.9 |  |
| Hospital location/teaching status, % |  |  | <0.001 |
| Rural location | 10.1 | 8.8 | 0.030 |
| Urban location | 23.1 | 17.6 |  |
| Urban teaching hospital | 66.9 | 73.6 |  |

LOS, length of hospital stay; THC, total hospital costs; SNF, skilled nursing facility; US\$, United States Dollar; BDD, body dysmorphic disorder
\* Significant at values <.05
\*\* The median income quartiles are defined as: first = \$1 - \$49,999; second = \$50,000 - \$64,999; third = \$65,000 - 85,999; and fourth = \$86,000 or more.

The study also assessed the burden of comorbidities in the BDD cohort. Specifically, 83.8% of individuals with BDD had a Charlson comorbidity index (CCI) score of 0, indicating less than 4% 10-year mortality risk. Approximately 12.9% of the BDD cohort had a CCI score of 1 (4% 10-year mortality risk). Among the non-psychiatric comorbidities observed in BDD hospitalizations, the most common were COPD (8.2%), diabetes (2.5%), and renal disease (1.4%).

Table 1 compares patient- and hospital-level variables for BDD hospitalizations with other hospitalizations for mental diseases and disorders.

### Indications for hospitalization

Out of a total of 3,384 BDD hospitalizations included in the study, 94.6% (3,201) had at least one other psychiatric comorbidity. The most common comorbidities included social anxiety disorder (40%), major depressive disorder (22.8%), bipolar II disorder (2.2%), substance use disorders (18.9%), schizoaffective disorder (1.5%), schizophrenia (1.5%), intentional self-harm (1.3%). The specific principal diagnoses are summarized in Table 2.

**Table 2.**
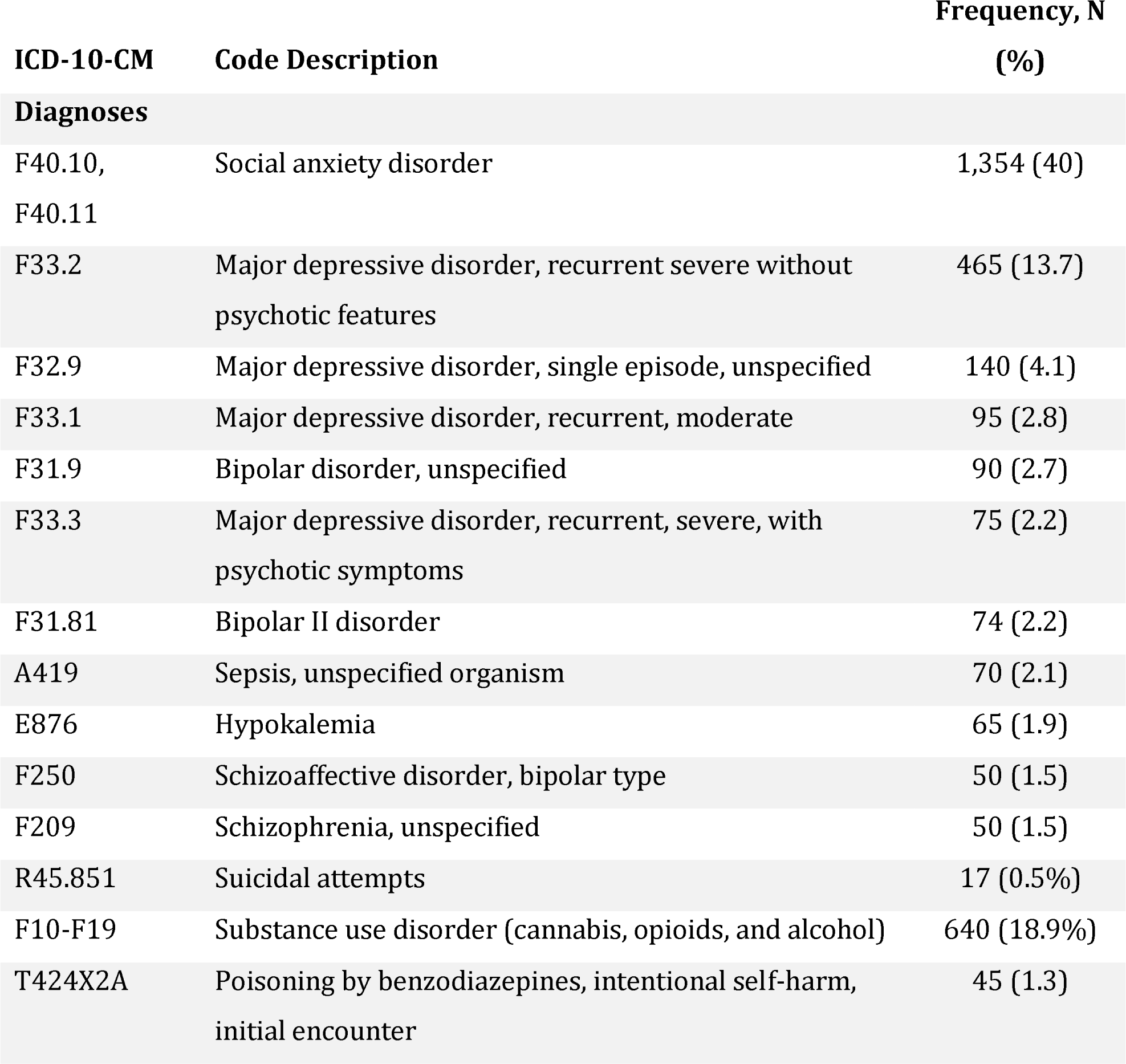
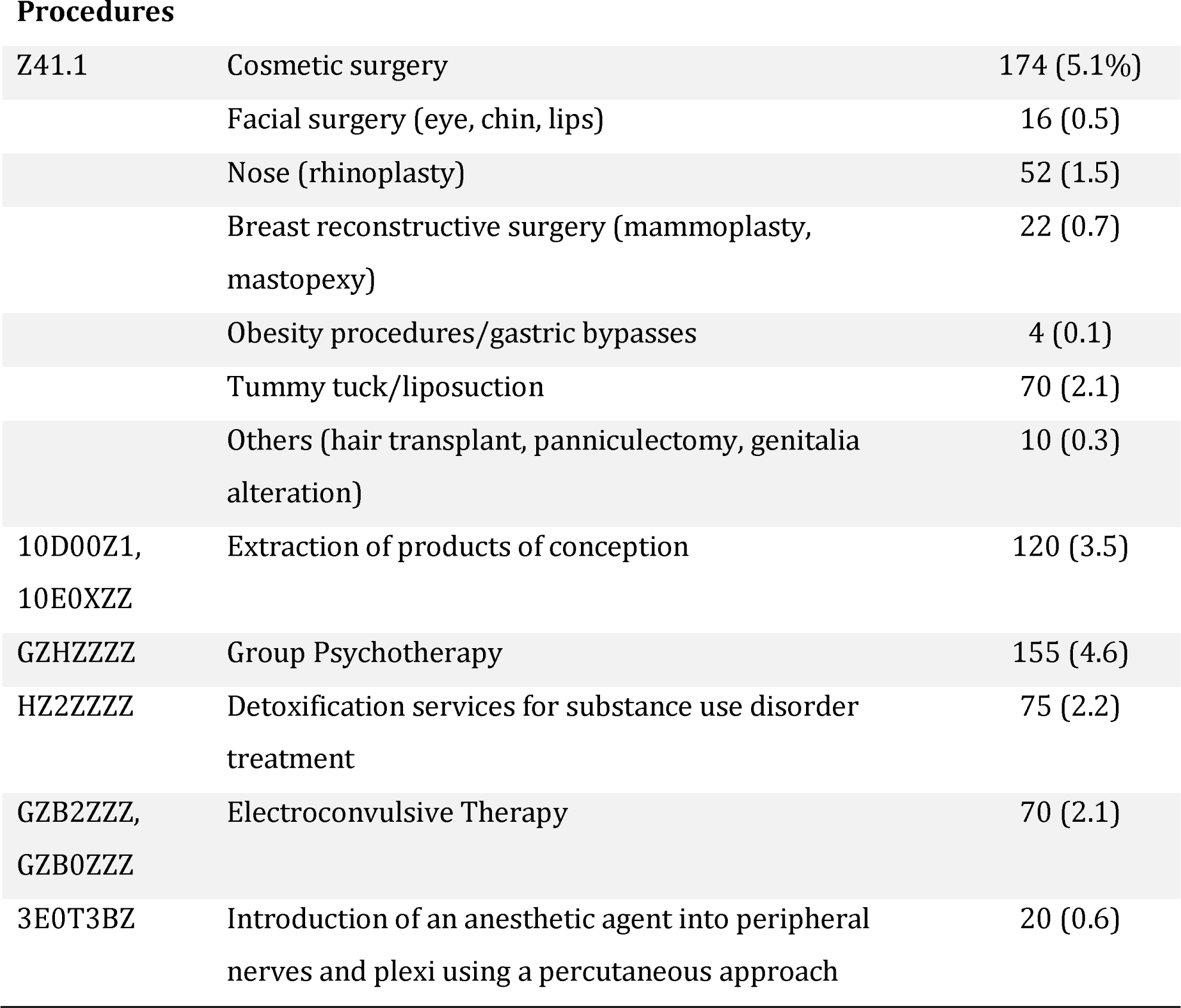
Most recurrent comorbidities and procedures associated with BDD hospitalizations.

| ICD-10-CM | Code Description | Frequency, N (%) |
| --- | --- | --- |
| **Diagnoses** |  |  |
| F40.10,<br>F40.11 | Social anxiety disorder | 1,354 (40) |
| F33.2 | Major depressive disorder, recurrent severe without psychotic features | 465 (13.7) |
| F32.9 | Major depressive disorder, single episode, unspecified | 140 (4.1) |
| F33.1 | Major depressive disorder, recurrent, moderate | 95 (2.8) |
| F31.9 | Bipolar disorder, unspecified | 90 (2.7) |
| F33.3 | Major depressive disorder, recurrent, severe, with psychotic symptoms | 75 (2.2) |
| F31.81 | Bipolar II disorder | 74 (2.2) |
| A419 | Sepsis, unspecified organism | 70 (2.1) |
| E876 | Hypokalemia | 65 (1.9) |
| F250 | Schizoaffective disorder, bipolar type | 50 (1.5) |
| F209 | Schizophrenia, unspecified | 50 (1.5) |
| R45.851 | Suicidal attempts | 17 (0.5%) |
| F10-F19 | Substance use disorder (cannabis, opioids, and alcohol) | 640 (18.9%) |
| T424X2A | Poisoning by benzodiazepines, intentional self-harm, initial encounter | 45 (1.3) |
| Procedures |  |  |
| Z41.1 | Cosmetic surgery | 174 (5.1%) |
|  | Facial surgery (eye, chin, lips) | 16 (0.5) |
|  | Nose (rhinoplasty) | 52 (1.5) |
|  | Breast reconstructive surgery (mammoplasty, mastopexy) | 22 (0.7) |
|  | Obesity procedures/gastric bypasses | 4 (0.1) |
|  | Tummy tuck/liposuction | 70 (2.1) |
|  | Others (hair transplant, panniculectomy, genitalia alteration) | 10 (0.3) |
| 10D00Z1,<br>10E0XZZ | Extraction of products of conception | 120 (3.5) |
| GZHZZZZ | Group Psychotherapy | 155 (4.6) |
| HZ2ZZZZ | Detoxification services for substance use disorder treatment | 75 (2.2) |
| GZB2ZZZ,<br>GZB0ZZZ | Electroconvulsive Therapy | 70 (2.1) |
| 3E0T3BZ | Introduction of an anesthetic agent into peripheral nerves and plexi using a percutaneous approach | 20 (0.6) |

The results indicated that 62.8% of these hospitalizations were associated with comorbidities of depression and anxiety. Suicidal attempts were reported in 0.5% of the cases, highlighting a concerning aspect of the disorder. Substance abuse, including cannabis, opioids, and alcohol, was observed in 18.9% of hospitalizations, indicating a significant co-occurrence with body dysmorphic disorder. In addition, a subset of patients (5.1%) underwent cosmetic surgery, with facial surgery (augmentation of the eyelids, chin or lips) accounting for 0.5%, nose surgery for 1.5%, and breast reconstructive surgery (mammoplasty, mastopexy) for 0.7%. Furthermore, 0.1% of patients received obesity procedures or gastric bypasses, whereas 2.1% underwent tummy tuck or liposuction procedures. A small percentage (0.3%) of patients underwent other cosmetic procedures, including hair transplantation, panniculectomy, and genitalia alteration.

### Hospitalization outcomes

Hospital stays for patients with BDD were, on average, a day longer than those for patients with other mental health conditions (P=0.002). A significant majority of BDD patients (89.8%) were routinely discharged to their homes, a rate higher than that in non-BDD cases (83.6%; P=0.008; Table 3). Transfers to skilled nursing facilities or intermediate care accounted for 5.8% of all discharges. Notably, there were no recorded instances of mortality among BDD hospitalizations, in contrast to a 0.1% (5,026) overall mortality rate in non-BDD-related hospitalizations. Additionally, the study noted 17 cases of suicide attempts among BDD patients, comprising 0.5% of the cases analyzed. BDD hospitalizations were associated with higher mean hospital costs than hospitalizations for other mental disorders combined ($34,314 vs. $27,770; P<0.001).

**Table 3.** Hospitalization outcomes for BDD compared with those for other mental health disorders.

| **Outcomes** | **Non-BDD hospitalizations**<br>**(N = 5,025,910), % unless**<br>**otherwise specified** | **BDD hospitalizations**<br>**(N= 3,384), % unless**<br>**otherwise specified** | **P-**<br>**value\*** |
| --- | --- | --- | --- |
| Discharge disposition |  |  | 0.008 |
| Routine home discharge | 83.6 | 89.8 |  |
| Transfer to a short-term<br>hospital | 2.2 | 1.7 |  |
| Other transfers (SNF or<br>intermediate care) | 10.8 | 5.8 |  |
| Home health care | 2.0 | 1.1 |  |
| Against medical advice | 1.4 | 1.7 |  |
| All-cause mortality | 0.09 | 0.0 |  |
| Mean THC, US\$ | 27,770 | 34,314 | <0.001 |
| Aggregate hospital<br>charges, US\$ | 139 billion | 61.8 million | 0.001 |
| Mean LOS | 7.8 days | 9 days | 0.002 |
| US\$, United States dollar; THC, total hospital charges; LOS, length of hospital stay; SNF, skilled nursing facility | | | |
| * Statistically significant at values <0.05 |  |  |  |

## DISCUSSION

In this study, we explored the hospitalization patterns among patients admitted for BDD. Typically, BDD onset occurs in adolescence, with symptoms intensifying to meet the full criteria of the Diagnostic and Statistical Manual of Mental Disorders, 5th edition (DSM-5) over time. Prior research indicates that BDD onset occurs before age 18 in two-thirds of cases, often around ages 12-13, suggesting a link with puberty and other physical and emotional changes.^17,18^ However, BDD can begin as early as age 4 and as late as the 40s.^18^ Early-onset BDD, diagnosed at age 17 or younger, tends to have a gradual onset, more severe symptoms, and a history of physical violence and psychiatric hospitalization.^19^

Our study found that the mean age of patients hospitalized with BDD was 34 ± 0.5 years, significantly younger than those admitted for other mental diseases (44 ± 0.1 years; P<0.001). Psychiatric hospitalization for BDD is primarily used for symptom stabilization during mental health crises, often necessitated by severe symptoms or risk to the patient’s safety. Criteria for psychiatric inpatient admission may be met when an individual with BDD is unable to keep up with daily responsibilities or poses immediate risk or danger to themselves.^20–22^

The findings of the index study shows that most hospitalizations among patients with BDD are for non-BDD-related diagnoses. Patients with BDD tend not to disclose their appearance concerns unless directly asked, leading to frequent underdiagnosis.^23^ The point prevalence of BDD in the general population is estimated at 2-3%, but it often goes unrecognized because of shame, embarrassment, or a desire for cosmetic rather than mental health treatment. Over 5% of BDD patients in the index study underwent cosmetic and weight reduction surgeries, despite BDD often being considered a contraindication for such treatments.^24^ BDD is typically seen as a contraindication to plastic surgery because of several reasons centered around the psychological nature of the disorder. Individuals with BDD often have unrealistic expectations of surgery, expecting it to completely resolve their distorted perceptions of physical flaws, which is rarely achievable. This dissatisfaction can lead to a cycle of persistent preoccupation, where the surgery fails to alleviate the patients’ obsessive concerns, sometimes shifting focus to other body parts or remaining dissatisfied with the operated area.^25^ Furthermore, undergoing surgery can exacerbate BDD symptoms, leading to heightened anxiety, depression, and even suicidal thoughts.^26^ A key challenge is the lack of insight in patients with BDD, who may not recognize their distress as stemming from a mental health disorder rather than a physical imperfection. This often results in postoperative dissatisfaction, repeated surgical procedures, and potential legal disputes with surgeons. Given these risks and the ethical implications of operating on individuals driven by a mental health disorder, the recommended approach is psychological interventions such as cognitive-behavioral therapy, which focuses on treating the underlying condition rather than the perceived physical imperfections.^27,28^

The findings of this study also corroborate the high prevalence of substance use disorders in patients with BDD. Suicidal attempts were reported in 0.5% of cases, echoing recent studies that link BDD with a substantial risk of suicidal ideation and behaviors, especially in late adolescence and early adulthood.^29^ Approximately 80% of individuals with BDD are reported to have experienced suicidal ideation, and 24%–28% have attempted suicide. The risk of suicidality in patients with BDD appears to be independent of comorbidity, although certain conditions, such as major depressive disorder and substance use disorders, may exacerbate it.^29^ In a nationwide epidemiologic study conducted in Germany, 31% of participants with BDD reported thoughts about committing suicide specifically because of appearance concerns, and 22% had actually attempted suicide.^30^ Recent data on completed suicide rates in individuals with BDD are limited, but older studies indicate that these rates are approximately 45 times higher than those in the general U.S. population.^31^ The relationship between BDD and suicidality is reportedly independent of comorbidity in an acute psychiatric setting.^32^ However, certain comorbid conditions may further exacerbate this relationship, such as comorbid major depressive disorder, posttraumatic stress disorder, and substance use disorder.

Our study found a higher prevalence of BDD in women (67.6%) than in men. BDD is more prevalent in women than in men, a trend influenced by several cultural, societal, and psychological factors. Women face heightened societal and cultural pressures regarding physical appearance, and they often internalize these rigorous beauty standards more than men. This societal emphasis on female beauty contributes to greater body image dissatisfaction and preoccupation with perceived physical imperfections. Women are more prone to comorbid mental health issues, such as depression and anxiety, which can exacerbate or coincide with BDD symptoms. Additionally, sex differences in reporting and seeking help for such disorders play a role; men may be less likely to acknowledge or seek treatment for BDD, possibly due to societal stigma or the perception that such concerns are less masculine. Traditional gender roles and socialization processes also contribute to this disparity, as women are often more attuned to their appearance and minor flaws, reflecting the pervasive focus on female appearance in media and societal norms. Although BDD is significantly impactful for all genders, these factors collectively contribute to its higher reported incidence in females.

### Strengths and Limitations

The NIS is a valuable resource for healthcare research, but it has limitations. As an administrative database, it relies on coded data, which can lead to inaccuracies due to coding errors or variations in coding practices across institutions. The NIS captures data from hospitalizations but excludes outpatient visits, thus providing a limited view of diseases and treatments that are primarily managed in outpatient settings. Additionally, the NIS contains limited information on patient follow-up, making it challenging to study long-term outcomes. It also lacks certain patient-specific details, such as lifestyle factors or severity of symptoms, which can be crucial for understanding disease progression and treatment effectiveness. The cross-sectional nature of the data limits the ability to establish causality, and because it is based on hospital discharges, not unique patients, there’s potential for double-counting patients with multiple admissions. Despite these limitations, the NIS remains a robust tool for understanding trends and patterns in inpatient care for BDD across the United States.

## CONCLUSION

Hospitalizations for body dysmorphic disorder, predominantly in young females, are marked by considerable psychiatric comorbidity, extended hospital stays, and elevated costs compared to other mental health conditions. Individuals with BDD often pursue cosmetic and surgical procedures to correct their perceived flaws. Therefore, early screening and diagnosis, both in outpatient settings and during hospital stays, are vital. This enables the initiation of prompt psychoeducation and brief interventions, followed by continued outpatient treatment, to effectively manage BDD symptoms.

### CRediT authorship contribution statement

OO participated in the conceptualization, design, investigation, data analysis, and writing of the manuscript.

KA participated in the conceptualization, design, investigation, data analysis, and writing of the manuscript.

SU participated in data curation, formal analysis, and writing of the original draft.

FU participated in the design, investigation, validation, methodology, and writing of the manuscript.

NO participated in the methodology, investigation and evaluation of the study.

CO participated in the design, formal analysis, and writing of the manuscript.

HY participated in the data analysis, writing-original draft, and writing-review and editing.

MO participated in the data curation, formal analysis, and writing-review and editing of the manuscript.

MB, VO, and NU participated in project supervision, validation, visualization and writing-review and editing of the manuscript.

### Consent for publication

All authors have read and approved the manuscript to be published.

## Data Availability

Publicly available National Inpatient Sample (NIS) datasets were utilized for this study. NIS data is available upon request from the Agency for Healthcare Research and Quality.

https://cdors.ahrq.gov/databases

## Acknowledgments

None

